# Modelling Quarantine Effects on SARS-CoV-2 Epidemiological Dynamics in Chilean Communes and their Relationship with the Social Priority Index

**DOI:** 10.1101/2022.01.21.22269658

**Authors:** Dino G. Salinas, María Leonor Bustamante, Mauricio O. Gallardo

## Abstract

An epidemiological model [Susceptible, Un-quarantined infected, Quarantined infected, Confirmed infected (SUQC)] has previously been developed and applied to incorporate quarantine measures and calculate COVID-19 contagion dynamics and pandemic control in some Chinese regions. Here, we generalized this model to incorporate the disease recovery rate and applied our model to records of the total number of confirmed cases of people infected with the SARS-CoV-2 virus in some Chilean communes. In each commune, two consecutive stages were considered: a stage without quarantine and an immediately subsequent quarantine stage imposed by the Ministry of Health. To adjust the model, typical epidemiological parameters were determined, such as the confirmation rate and the quarantine rate. The latter allowed us to calculate the reproduction number. The mathematical model adequately reproduced the data, indicating a higher quarantine rate when quarantine was imposed by the health authority, with a corresponding decrease in the reproduction number of the virus down to values that prevent or decrease its exponential spread. In general, during this second stage, the communes with the lowest social priority indices had the highest quarantine rates, and therefore, the lowest effective viral reproduction numbers. This study provides useful evidence to address the health inequity of pandemics. The mathematical model applied here can be used in other regions or easily modified for other cases of infectious disease control by quarantine.

## 1. INTRODUCTION

Mathematical models of the spread of infectious diseases can be very useful for monitoring and controlling epidemics when such models use and predict the number of infected people. These figures are frequently reported by health authorities during a pandemic, as in the current coronavirus (COVID-19) pandemic, which is caused by the severe acute respiratory syndrome coronavirus 2 (SARS-CoV-2)[1]. In many countries, periodic reports on the total number of confirmed cases by region have been issued, and these reports have helped the population follow the pandemic dynamics. However, unlike the model that we propose and apply here, not all mathematical models of academic interest meet the practical requirement of fitting the data included in official epidemiological reports.

Dynamic models, such as Susceptible-Infectious-Removed (SIR), Susceptible-Exposed-Infectious-Removed (SEIR), and others [2-4], are difficult to apply to SARS-CoV-2 because of three main characteristics: the relatively long incubation period, artificial factors (medical resources and quarantines), and variations in the efficiency of confirmation methods. For this reason, an appropriate model for these characteristics in our study of SARS-CoV-2 propagation dynamics could be the SUQC model [5], which was applied to data of the total number of daily confirmed cases recorded in official Chinese reports from January and February 2020. Using this model, it was determined the values of three epidemiological parameters useful for epidemic control, monitoring, intervention, and evaluation: the reproduction number [6, 7], quarantine rate, and confirmation rate. Below, we review this model and then propose its generalization and subsequent application to some Chilean communes.

For the SUQC model [5], and assuming a commune of *N* inhabitants, the following types of individuals were considered: *S*, susceptible to infection; *U*, infected, unquarantined, infectious (unlike *E* in the SEIR model) and either presymptomatic or symptomatic; *Q*, infected and quarantined, and therefore, non-infectious, deriving from a *U*, hospitalized or isolated; and *C*, confirmed infected case. The *R* of the SIR and SEIR models is disregarded in SUQC model, which assumes there is no recovery or death. The total number of individuals of type *S, U, Q*, and *C* at time *t* is indicated as *S*_*t*_, *U*_*t*_, *Q*_*t*_, and *C*_*t*_, respectively.

The epidemiological parameters of the SUQC model are as follows: *α*, the number of individuals infected by an unquarantined individual per day – the infection rate 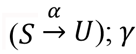, the quarantine rate 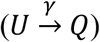; and *β*, the probability that *Q* is confirmed as *C*, i.e., the confirmation rate 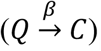.

The model is defined by the following system of ordinary differential equations:

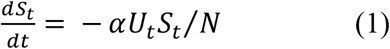

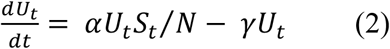

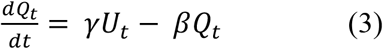

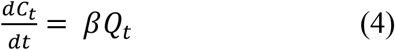

Note that, according to Eq. 4, *C*_*t*_ corresponds to the total number of cases of infection confirmed until time *t* (i.e., the cumulative number of cases).

Proposing a generalization of the previous model, we have modeled the spread of SARS-CoV-2 in some Chilean communes of the Metropolitan Region (including Santiago, the capital of the country), over the period of a few months during 2020, under different quarantine measures, depending on whether they were spontaneous or imposed by the health authority. The generalized model proposed by us, SUQCR, unlike the SUQC model, includes recovered individuals (*R*) depending on the recovery rate of infectious individuals (*U* and *Q*). Dead individuals are not included in the model. In addition to proposing the SUQCR model, this study aimed to demonstrate its applicability, calculating and comparing epidemiologically relevant fitting parameters of each commune according to its quarantine measures. It also aimed to assess whether these results have any correlation with the Social Priority Index (SPI), a parameter that indicates the social priority in each commune studied [8]. In this regard, the most vulnerable populations have endured a greater impact of COVID-19 worldwide [9], according to sociodemographic factors such as access to healthcare [10]. Accordingly, we believe that this study contributes new scientific evidence for addressing health inequity during a pandemic.

## 2. METHODOLOGY

### 2.1. Definition of the SUQCR model

We define the SUQCR model by including recovered individuals (*R*) in the SUQC model. *R* represents individuals who are no longer infectious, who are considered recovered, and who derive from *Q* or *U*, at a rate of 1/ *D*; as such, the mean time of recovery from the disease is *D*, in days. The factor 1/ *D* does not apply to individuals *C* because they correspond to cases counted as confirmed at any time up to *t*. Thus, the SUQCR model that we propose here as a generalization of the SUQC model is defined in the scheme of Figure 1 and by the following system of ordinary differential equations:

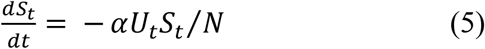

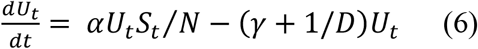

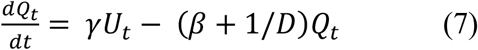

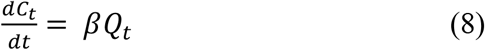

To demonstrate the relationship between the SUQC and SUQCR models, note that Eqs. 5 and 8 are the same as Eqs. 1 and 4, respectively. In particular, Eqs. 6 and 7 converge to Eqs. 2 and 3. respectively, when *D* ≫ 1. This demonstrates that SUQCR is a generalization of SUQC.

**Figure 1.**
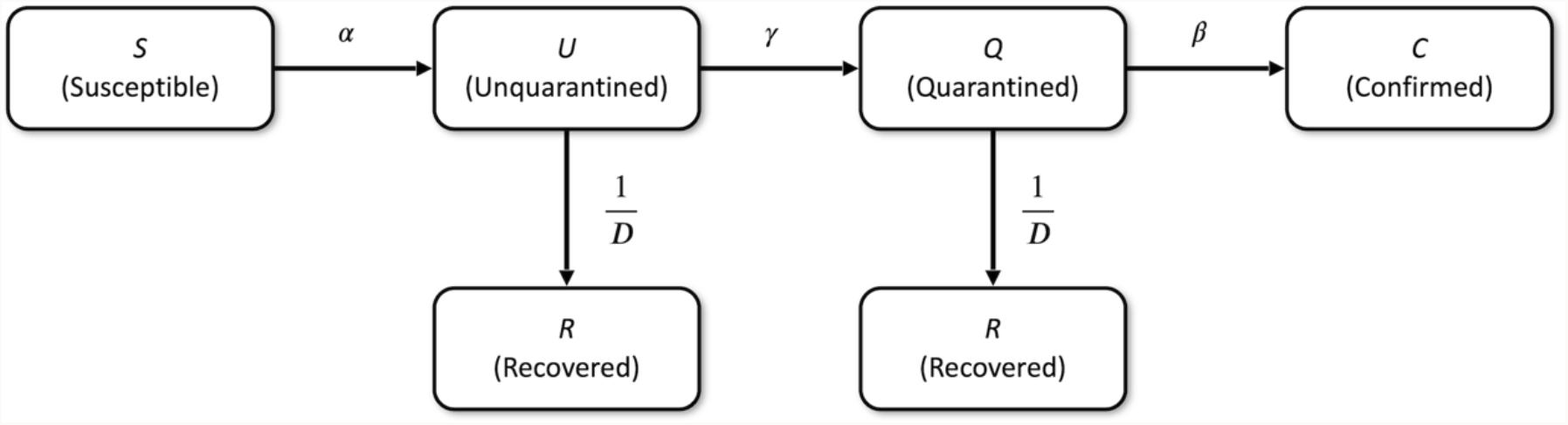
Schematic of the SUQCR model. The proposed model in this study (Eqs. 5-8) is a generalization of the SUQC model [5], which includes *R* individuals, who are no longer infectious, are considered recovered, and derive from *Q* or *U*, at a rate of 1/ *D*.

### 2.2. Study communes

The modeled data were the values of *C*_*t*_ recorded in each of the 32 communes of the province of Santiago, in the Metropolitan Region of Chile. The data from each of these communes were grouped into two consecutive stages. The first stage (stage 1) is without quarantine imposed by the health authorities. Given the influence of national and international news about the pandemic, the constant advice of subject-matter experts and quarantines previously imposed in some communes of the country, a fraction of the population most likely followed some degree of quarantine and other social distancing measures in stage 1, in addition to hygiene protocols and the use of a masks [11], albeit voluntarily. In the second stage (stage 2), quarantine was officially imposed by the health authorities, mandating compliance with strict social distancing protocols and other self-care practices.

### 2.3. Total number of confirmed cases of infection (*C*_*t*_) by commune during the study period

The values of *C*_*t*_ for each of the 32 communes are from the epidemiological records of the period (corresponding to stages 1 and 2 for all communes) spanned from March 30 to June 15, 2020. [12].

### 2.4. Calculation of epidemiological parameters of the SUQCR model

The value of *α* could no longer be measured in pure form through an exponential model simpler than the model that we applied here because most of the population already had enough information to adopt some self-care measures by the time the pandemic arrived in Chile. However, such a simple exponential approximation was used in China with the first data collected in Wuhan, where the first-ever cases were recorded. As a result, the value of the infection rate, *α* = 0.2967, calculated for Wuhan from January 20 to 27, 2020, was used in the SUQC model, as applied to a few other regions of China. In this model, the reproduction number, ℛ, was calculated considering that fluctuations in *α* could be compensated by fluctuations in *γ* because ℛ = *α*/*γ* [5]. However, when *γ*=0 this ℛ value is indeterminate. In our SUQCR model, we considered the aforementioned value of *α*, although ℛ should be calculated as follows:

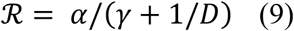

with *D* = 14 days [13]. Although during the study period the number of susceptible individuals almost does not decrease compared to the total population (*S*_*t*_ ≈ *N*), we consider ℛ as an effective reproduction number because ℛ < ℛ_*basic*_ ≡ *αD*.

Each quarantine stage, 1 and 2 (i.e., voluntary and imposed), has the following time series: a time series of *C*_*t*_, denoted by **C**, derived from the study period, and a theoretical time series, denoted by **Ĉ**, corresponding to the same days, determined by numerical integration of the Eqs. 5-8 system using the Runge-Kutta method and described as a function *f*() of the fitting parameters.

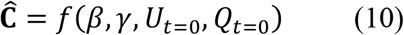

The fitting parameters *β, γ, U*_*t*=0_, and *Q*_*t*=0_ were calculated by minimizing the error function *err*, which is equal to the mean squared deviation between the empirical and theoretical time series data:

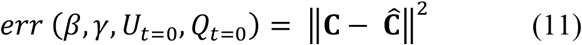

In fitting the parameters, to minimize *err* (Eq. 11), we alternatively used the MATLAB function *fminsearch* or *fmincon* [14] to optimize the fit of the model.

The data and programs used are available as supplementary material.

### 2.5. Study of the estimated parameters by commune

After applying the SUQCR model to each commune and in each quarantine stage, the goodness-of-fit parameters were recorded, and the values of *β, γ* and R_effective_ were analyzed for stages 1 and 2. Particularly the correlations between *γ* and SPI were studied. The SPI is used by the Metropolitan Regional Government to support the communes with the greatest relative deficiencies (Table 2). This index is a composite indicator that integrates relevant aspects of social development (income, education, and health), and its numerical value allows us to measure the relative standard of living of the population of a commune.

## 3. RESULTS

We applied the SUQCR mathematical model to data of total confirmed cases of infection during approximately three months, in stages 1 and 2, for each of the 32 study communes, thereby calculating the Pearson’s correlation coefficient, *r*, and the corresponding fitting parameters for each commune. As a case in point, Table 1 outlines the parameters of the SUQCR model for 3 communes, and Figure 2 shows a typical curve fitting of the model to the total number of confirmed cases reported in a commune.

**Table 1.** Goodness-of-fit results of some epidemiologically relevant parameters of the three Chilean communes studied. As defined by Eqs. 5-8, the model was fitted to the records of the number of total confirmed infected cases (*C*_*t*_), which were sourced from the Ministry of Health. The start and end dates of each stage are indicated.

|  |  | San Ramón<br>(Population: 86,510) |  | Recoleta<br>(Population: 190,075) |  | Santiago Centro<br>(Population: 503,147) |  |
| --- | --- | --- | --- | --- | --- | --- | --- |
|  |  | Stage I<br>(30-March-2020 to 8-May-2020) | Stage II<br>(11-May-2020 to 15-June-2020) | Stage I<br>(30-March-2020 to 5-May-2020) | Stage II<br>(8-May-2020 to 15-June-2020) | Stage I<br>(30-March-2020 to 4-May-2020) | Stage II<br>(8-May-2020 to 15-June-2020) |
| $\alpha$ [day<sup>-1</sup>] | [5] | 0.2967 | 0.2967 | 0.2967 | 0.2967 | 0.2967 | 0.2967 |
| $D$ [day] | [13] | 14 | 14 | 14 | 14 | 14 | 14 |
| $U_{t=0}$ | Calculated parameter | 1.7708 | 227.6755 | 12.8968 | 470.0697 | 90.9906 | 1795.5073 |
| $Q_{t=0}$ | Calculated parameter | 22.9903 | 295.4115 | 0.6689 | 110.4116 | 0.0000 | 154.6483 |
| $\beta$ [day<sup>-1</sup>] | Calculated parameter | 0.4208 | 0.6370 | 0.6410 | 0.6102 | 0.3047 | 0.1096 |
| $\gamma$ [day<sup>-1</sup>] | Calculated parameter | 0.0391 | 0.2019 | 0.1060 | 0.2042 | 0.1740 | 0.2220 |
| $\mathcal{R}$ | Eq. 9 | 2.6832 | 1.0856 | 1.6723 | 1.0763 | 1.2091 | 1.0113 |
| $r^2$ | Square of the Pearson's correlation coefficient | 0.9820 | 0.9912 | 0.9836 | 0.9909 | 0.9762 | 0.9990 |

**Table 2.** Goodness-of-fit of the parameters assessed for the SUQCR model (Eqs. 5-8). In total, 32 communes were studied in the province of Santiago, Chile. The continuous interval of days corresponding to stages 1 and 2 was extended for all communes from March 30 to June 15, 2020. The number of inhabitants (*N*) and the social priority index (SPI) are also shown for each commune. Values of SPI are for communes in 2020, available as IPS 2020 in the report of Regional Ministerial Secretariat for Social Development and Metropolitan Family (*Secretaría Regional Ministerial de Desarrollo Social y Familia Metropolitana*) [8].

| City | $N$ | SPI | Stage 1 | | | | Stage 2 | | | |
| --- | --- | --- | --- | --- | --- | --- | --- | --- | --- | --- |
| | | | $\gamma$<br>[day<sup>-1</sup>] | $\beta$<br>[day<sup>-1</sup>] | $\mathcal{R}$ | $r^2$ | $\gamma$<br>[day<sup>-1</sup>] | $\beta$<br>[day<sup>-1</sup>] | $\mathcal{R}$ | $r^2$ |
| Lo Prado | 104,403 | 77.71 | 0.1512 | 0.8971 | 1.3328 | 0.9937 | 0.1611 | 0.3124 | 1.2758 | 0.9942 |
| Quinta Normal | 136,368 | 73.17 | 0.1385 | 0.4473 | 1.4133 | 0.9953 | 0.1672 | 0.8299 | 1.2435 | 0.9983 |
| La Granja | 122,557 | 76.37 | 0.1281 | 0.7785 | 1.4873 | 0.9914 | 0.1684 | 0.3836 | 1.2372 | 0.9526 |
| Renca | 160,847 | 72.85 | 0.1321 | 0.4428 | 1.4581 | 0.9967 | 0.1693 | 0.7057 | 1.2328 | 0.9952 |
| Estación Central | 206,792 | 71.67 | 0.1569 | 0.9234 | 1.2995 | 0.9959 | 0.1696 | 0.3635 | 1.2310 | 0.9777 |
| Cerro Navia | 142,465 | 85.91 | 0.1598 | 0.3463 | 1.2830 | 0.9976 | 0.1730 | 0.4011 | 1.2139 | 0.9958 |
| El Bosque | 172,000 | 80.97 | 0.1301 | 0.2469 | 1.4723 | 0.9852 | 0.1742 | 0.9965 | 1.2078 | 0.9984 |
| Independencia | 142,065 | 72.2 | 0.1006 | 0.5141 | 1.7244 | 0.9983 | 0.1824 | 0.9072 | 1.1689 | 0.9901 |
| La Pintana | 189,335 | 89.29 | 0.1415 | 0.5240 | 1.3935 | 0.9980 | 0.1829 | 0.2711 | 1.1668 | 0.9991 |
| Pudahuel | 253,139 | 67.64 | 0.1401 | 0.2495 | 1.4027 | 0.9986 | 0.1840 | 0.2644 | 1.1614 | 0.9942 |
| Pedro Aguirre Cerda | 107,803 | 75.65 | 0.1152 | 0.9151 | 1.5901 | 0.9567 | 0.1871 | 0.0545 | 1.1478 | 0.9967 |
| Cerrillos | 88,956 | 67.81 | 0.0476 | 0.4451 | 2.4917 | 0.9598 | 0.1877 | 0.4365 | 1.1448 | 0.9951 |
| La Cisterna | 100,434 | 70.21 | 0.1398 | 0.9053 | 1.4049 | 0.9919 | 0.1922 | 0.4830 | 1.1254 | 0.9970 |
| Lo Espejo | 103,865 | 88.83 | 0.1660 | 1.0000 | 1.2494 | 0.9902 | 0.1954 | 0.5875 | 1.1120 | 0.9974 |
| Peñalolén | 266,798 | 66.19 | 0.0488 | 0.8321 | 2.4680 | 0.9255 | 0.1993 | 0.4479 | 1.0958 | 0.9980 |
| Ñuñoa | 250,192 | 40.96 | 0.0950 | 0.7007 | 1.7830 | 0.9162 | 0.1997 | 0.5543 | 1.0942 | 0.9993 |
| Maipú | 578,605 | 60.86 | 0.1711 | 0.6727 | 1.2232 | 0.9822 | 0.2013 | 0.5732 | 1.0879 | 0.9989 |
| San Miguel | 133,059 | 56.63 | 0.1524 | 0.4472 | 1.3255 | 0.9911 | 0.2018 | 0.3575 | 1.0859 | 0.9987 |
| San Ramón | 86,510 | 83.5 | 0.0391 | 0.4208 | 2.6832 | 0.9820 | 0.2019 | 0.6370 | 1.0856 | 0.9912 |
| Conchalí | 139,195 | 79.87 | 0.1197 | 0.4339 | 1.5525 | 0.9931 | 0.2029 | 0.9958 | 1.0817 | 0.9936 |
| Recoleta | 190,075 | 76.6 | 0.1060 | 0.6410 | 1.6723 | 0.9836 | 0.2042 | 0.6102 | 1.0763 | 0.9909 |
| Macul | 134,635 | 57.63 | 0.1217 | 0.3818 | 1.5366 | 0.9955 | 0.2053 | 0.5144 | 1.0721 | 0.9995 |
| La Reina | 100,252 | 38.35 | 0.1575 | 0.2767 | 1.2963 | 0.9944 | 0.2083 | 0.4125 | 1.0608 | 0.9985 |
| La Florida | 402,433 | 64.22 | 0.1637 | 0.1078 | 1.2616 | 0.9823 | 0.2088 | 0.2390 | 1.0588 | 0.9992 |
| San Joaquín | 103,485 | 76.87 | 0.1373 | 0.1117 | 1.4212 | 0.9891 | 0.2119 | 0.4355 | 1.0471 | 0.9958 |
| Las Condes | 330,759 | 11.64 | 0.1805 | 0.5566 | 1.1775 | 0.9376 | 0.2160 | 0.4729 | 1.0324 | 0.9987 |
| Quilicura | 254,694 | 58.69 | 0.0323 | 0.8372 | 2.8605 | 0.8521 | 0.2177 | 0.3183 | 1.0262 | 0.9976 |
| Santiago Centro | 503,147 | 55.2 | 0.1740 | 0.3047 | 1.2091 | 0.9762 | 0.2220 | 0.1096 | 1.0113 | 0.9990 |
| Lo Barnechea | 124,076 | 35.08 | 0.1512 | 0.4810 | 1.3327 | 0.9937 | 0.2256 | 0.4664 | 0.9989 | 0.9998 |
| Huechuraba | 112,528 | 56.6 | 0.1439 | 0.9918 | 1.3782 | 0.9896 | 0.2279 | 0.4051 | 0.9911 | 0.9970 |
| Providencia | 157,749 | 24.91 | 0.1607 | 0.4301 | 1.2781 | 0.9864 | 0.2316 | 0.3368 | 0.9790 | 0.9997 |
| Vitacura | 96,774 | 7.94 | 0.1249 | 0.5546 | 1.5115 | 0.9812 | 0.2386 | 0.4164 | 0.9571 | 0.9980 |

**Figure 2.**
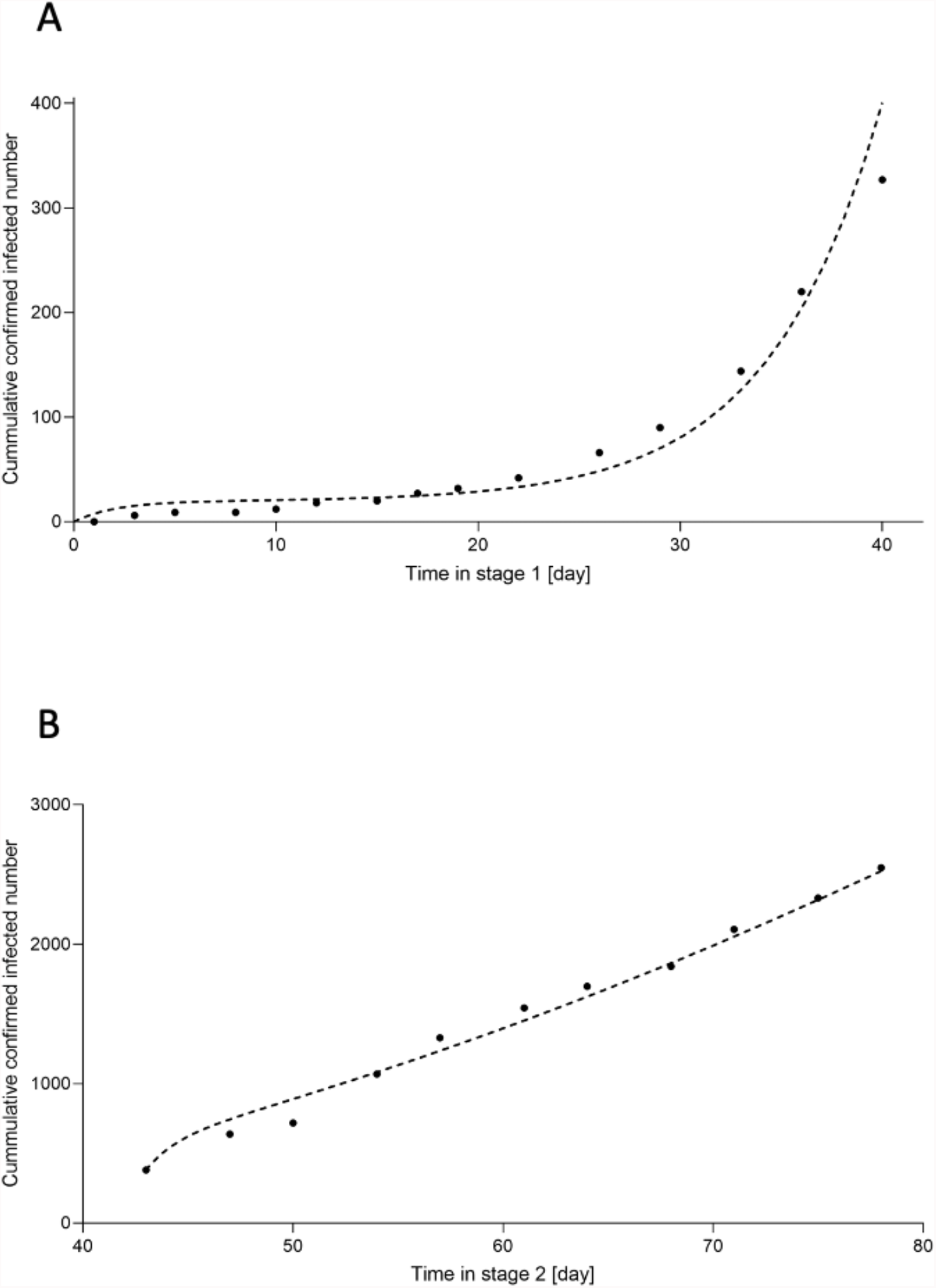
Curve fitting of the SUQCR model of the San Ramón commune. The data of total confirmed cases of the San Ramón commune, in the Metropolitan Region of Chile, were used. The goodness-of-fit results are outlined in Table 1. A) Without quarantine imposed by the health authorities (stage 1). B) With quarantine imposed by the health authorities (stage 2). The vertical legend corresponds to the total number of confirmed cases of infection (*C*_*t*_)

Table 2 outlines epidemiologically relevant parameters for the 32 communes in stages 1 and 2. There were 63 goodness-of-fit with *r*^2^ > 0.90, and one with *r*^2^ ≈ 0.85. In all communes, the ℛ decreased from stage 1 to stage 2.

Figure 3 shows a significant increase in mean *γ*, and thus, a significant decrease in mean ℛ from stage 1 to 2. In contrast, the mean *β* showed no significant change between stages.

**Figure 3.**
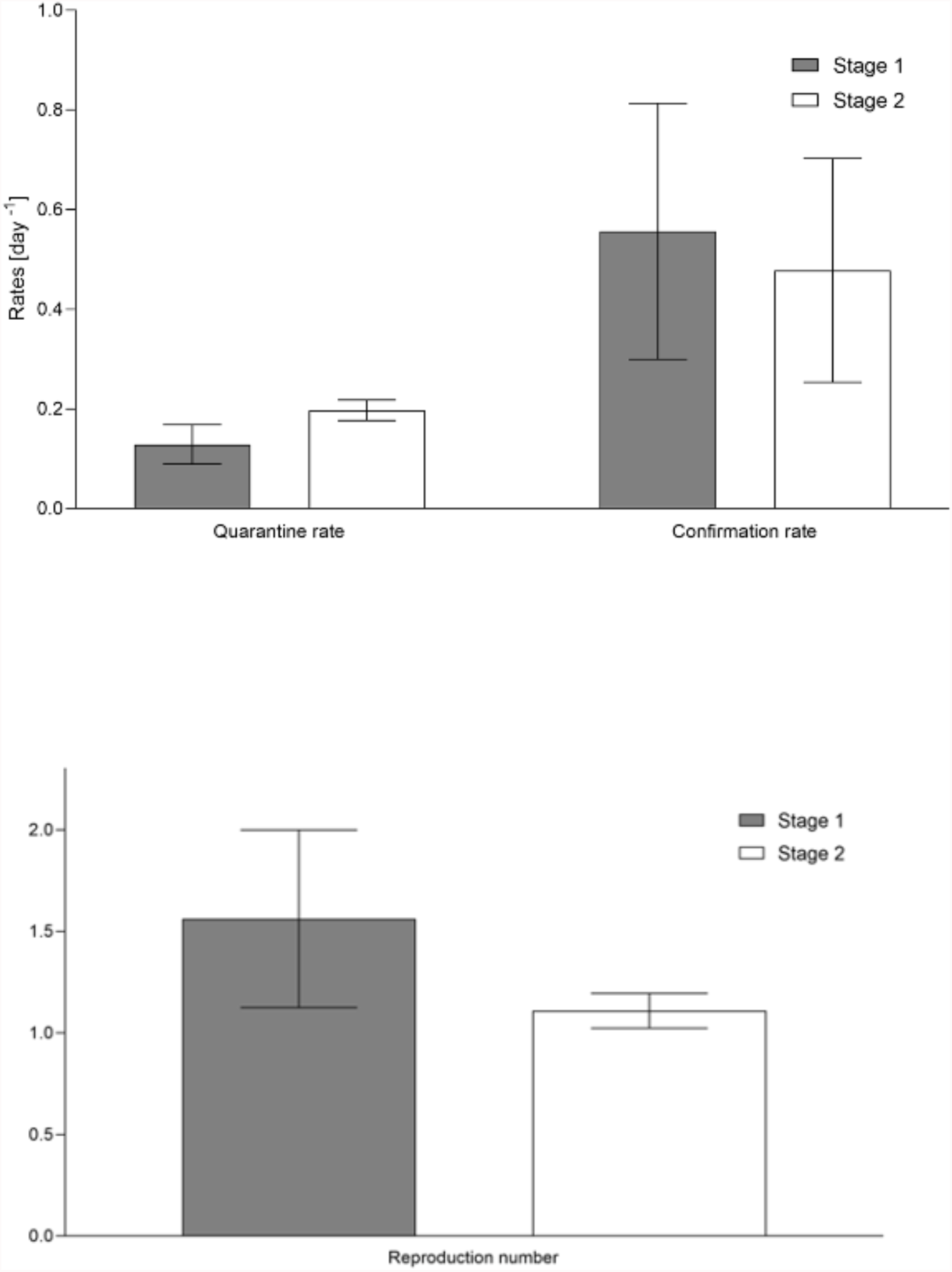
Comparison of mean goodness-of-fit of epidemiologically relevant parameters. The mean values per quarantine stage of the goodness-of-fit of the 32 communes of the province of Santiago, Chile, are compared for stages 1 and 2: quarantine rate (*γ*), confirmation rate (*β*) and reproduction number (ℛ, obtained from *γ* in Eq. 9). Student’s *t*-test was applied to the mean values of two paired samples, in two-tailed tests, assessing *p* < 0.01 for *γ* and ℛ, and *p* = 0.23 for *β*. The values of the parameters used for calculations are outlined in Table 2.

Figure 4 shows the ℛ values for each of the 32 communes in stages 1 and 2 (Table 2). From Figure 4 and Table 2, in the stage 2 and in qualitative terms, the communes with the 15 highest ℛ values had medium or high social priority (64.22 ≤ SPI ≤ 89.29, 1.2758 ≥ ℛ ≥ 1.0958). While the 7 lowest ℛ values were from communes without or with low social priority (7.94 ≤ SPI < 64.22, 0.9571 ≤ ℛ ≤1.0324). For the narrow intermediate range of ℛ (1.0324 < ℛ < 1.0958), corresponding to the 10 remaining communes, the social priority was of any kind.

**Figure 4.**
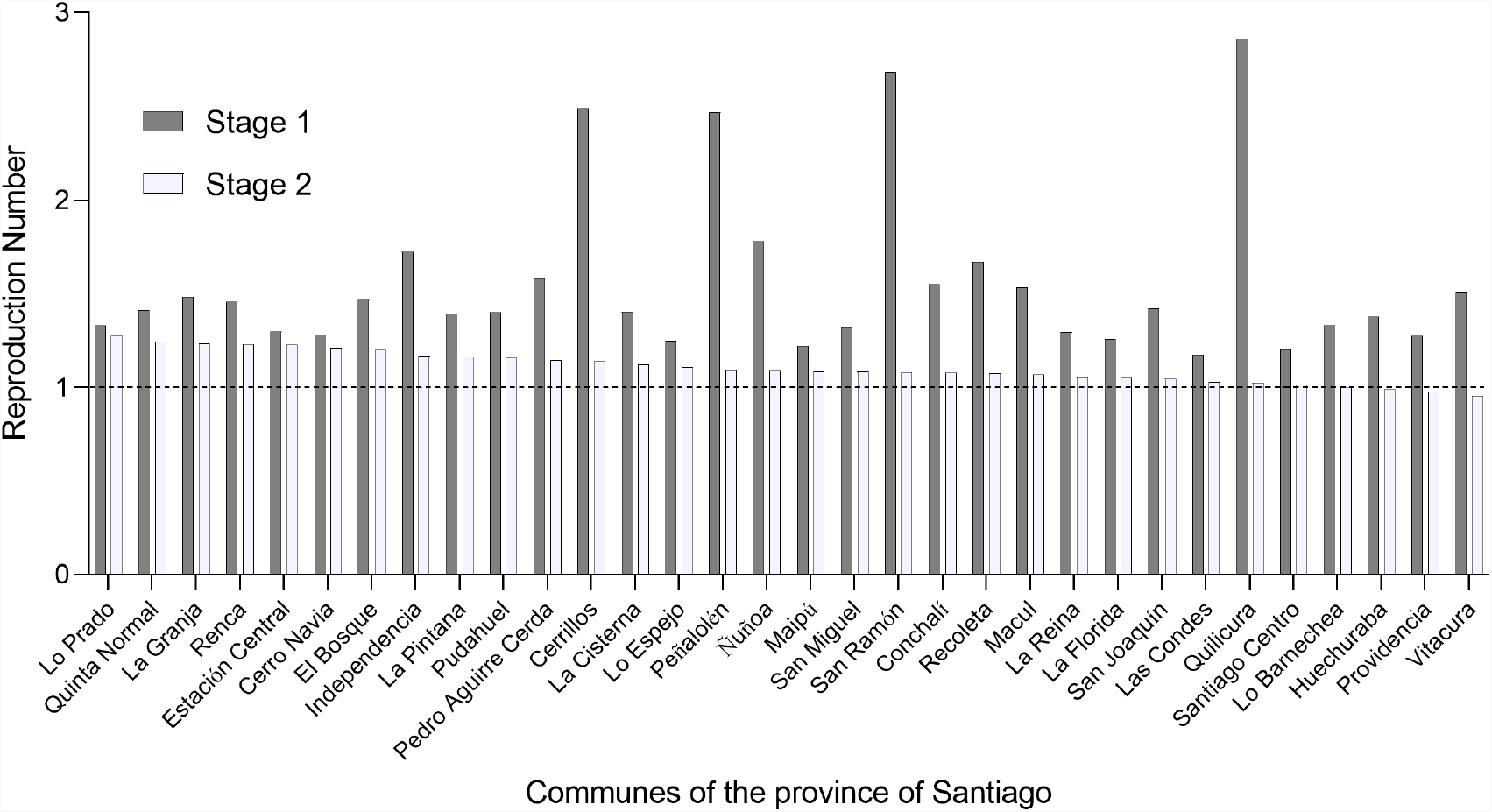
Comparison of reproduction numbers. The values of the reproduction number (ℛ) are compared in each quarantine stage of the 32 communes of the province of Santiago, in the Metropolitan Region of Chile. The values of the individual parameters are outlined in Table 2.

For the correlation between *γ* versus SPI in stages 1 and 2, using the values outlined in Table 2, *r*^2^ was 0.0360 and 0.4799, respectively (Figure 5).

**Figure 5.**
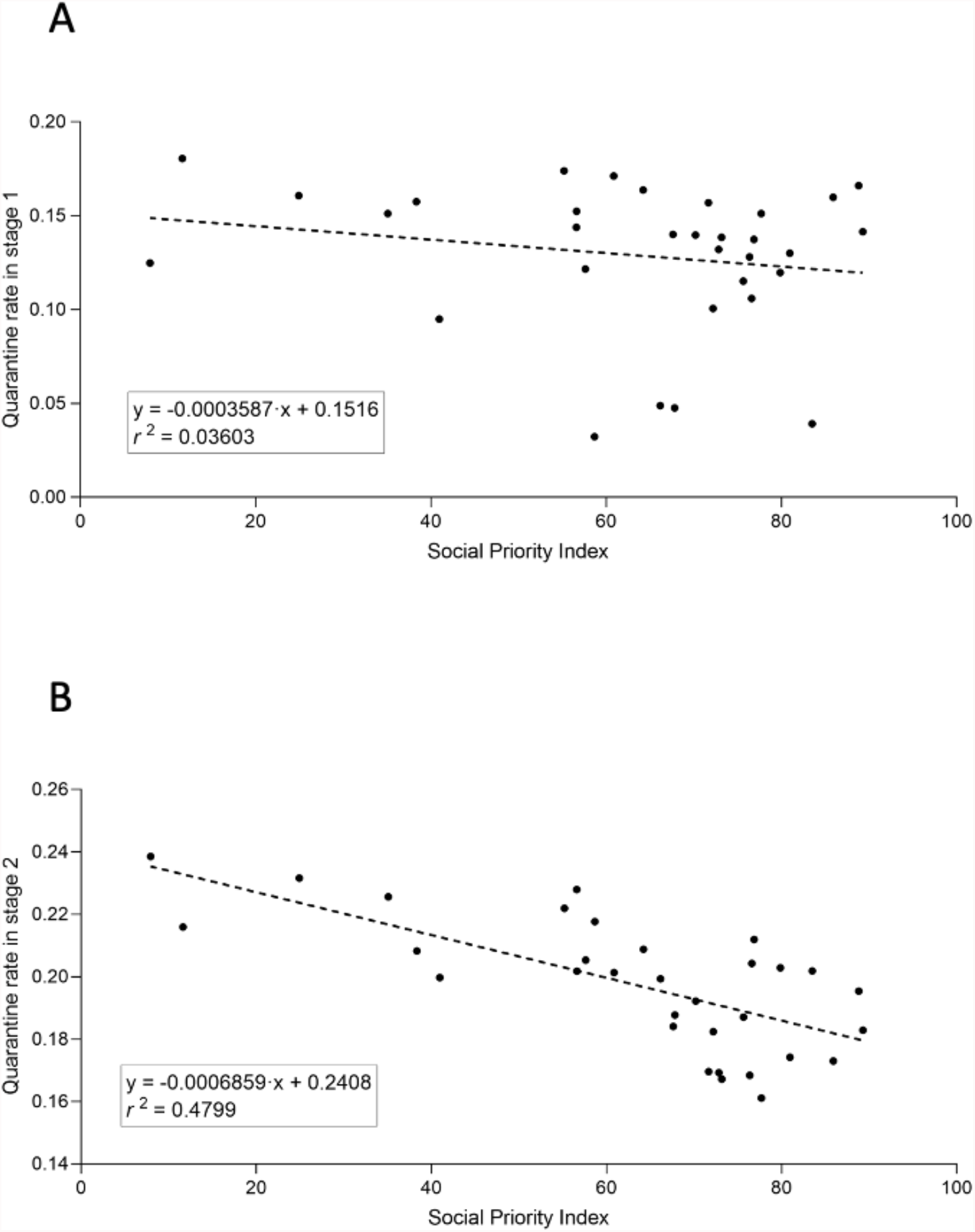
Reproduction number versus social priority index. The graphs indicate the regression equations and the square of the Pearson’s correlation coefficient (*r*^2^) between the quarantine rate (***γ*** [day ^-1^]) and the social priority index (SPI for communes in 2020) of the study communes. A) Without quarantine imposed by the health authorities (stage 1). B) With quarantine imposed by the health authorities (stage 2). The values of the parameters are outlined in Table 2.

## 4. DISCUSSION

Chile is currently one of the countries with the greatest control over the COVID-19 pandemic in terms of spread, virulence, and lethality indicators associated with SARS-CoV-2 and its genomic variants [15-18]. An important finding from the Chilean experience has been that, when localized lockdowns are applied, coordination across interconnected geographic areas is necessary to reverse the growth of disease transmission produced by indirect effects from neighboring areas [19]. In this regard, the lack of people flux between communes in our model could affect some results.

Because the effects of vaccination were not included in the mathematical model used in this study, we selected data from a period before COVID-19 vaccination, in an early stage of the pandemic in Chile. The study period was the same for all communes, with the only possible differences being the starting dates of the quarantine stages imposed by the health authorities (i.e., stage 2). In turn, all the communes chosen for this study are from the same region, and therefore, are highly homogeneous, with respect to climatic conditions, with their main differences in socio-demographic parameters, such as quality of life and access to healthcare.

In this study, we extended a known mathematical model of SARS-CoV-2 spread (SUQC model, Eqs. 1-4)[5], proposing a model that considers recovered patients, and therefore non-infectious (SUQCR model, Eqs. 5-8). We applied this model to determine key parameters for analysis, planning, and evaluation of social distancing and quarantine measures in the current COVID-19 pandemic. The modeling data used was the time series of the total number of confirmed cases of infection with SARS-CoV-2 from each commune of the province of Santiago, Chile. For this purpose, in each commune, two consecutive social distancing and quarantine stages were considered: the first was spontaneous, not officially mandated, and the second was the quarantine imposed by the health authorities, resulting in the typical goodness-of-fit and epidemiologically relevant parameters in each stage and commune. Of all the fitting parameters calculated in this study, we were especially interested in *γ*, owing to its utility in calculating ℛ, which also depended on *α* and *D*, both constant terms in this study. Significant differences were found in the mean of *γ* (increasing) and the mean of ℛ (consequently decreasing, by Eq. 9) between the two quarantine stages. Instead, no significant differences were found in the mean of *β* between the same two quarantine stages, in line with the maintenance of protocols and with the detection techniques for infected patients.

The quarantine measures imposed by the health authorities increased *γ* in all the communes studied. The increases in *γ* and the corresponding decreases in ℛ occurred in the first days of the quarantines in the stage 2, with characteristic values for the entire period. We believe that the study communes were already applying widely publicized self-care health recommendations during stage 1, such as avoiding crowds and closed environments, wearing a mask, and hand washing. The initial news about the beginning of the COVID-19 pandemic emerged from China in January 2020, and the first cases of COVID-19 in Chile were detected in early March 2020, when some more extensive health measures were implemented in airports and in some communes. In turn, assuming that all study communes made similar efforts to comply with the imposed quarantines (stages 2) should facilitate the interpretation of differences in their results during this stage. Accordingly, the differences between the values of ℛ in stage 2 likely resulted from structural, environmental, and social deficiencies in each commune.

Furthering the above, each commune is characterized by a set of environmental, cultural, economic, demographic, and population (mental and physical) health variables, which could influence the potential to comply with the social distancing or quarantine measures imposed by the health authorities in stage 2 and by the effectiveness of its compliance. As such, the relationship between SPI of a commune, an indicator of its socioeconomic deficiencies, and ℛ is interesting. Our results showed that under imposed quarantine, *γ* tends to increase as SPI decreases in the communes. In fact, the negative correlation between the *γ* and the SPI was stronger in stage 2 than in stage 1 (with *r*^2^ values of 0.4799 and 0.0360, respectively). In stage 2, communes without or with low social priority tended to avoid exponential growth more efficiently (ℛ ≈1 or ℛ < 1) than the other communes. These findings confirm that worse socioeconomic conditions make it more difficult to comply with or decrease the effectiveness of the pandemic control measures, such as social distancing and quarantines [20]. The correlation between *γ* and SPI was much weaker in stage 1, possibly due to the lack of imposed quarantine and self-care behavior of the population of the communes, which was rather spontaneous.

More precise or similar relationships could be calculated using a more extensive and varied database of epidemiological parameters. Accordingly, different countries should have epidemiological models similar to each other and share their databases on the relevant characteristics in each commune or region for increasing social distancing and compliance with the quarantine. These measures are the main non-invasive interventions that have proved useful in reducing the spread of COVID-19. In fact, the effect of the imposed lockdown measures in the communes appeared noticeably early. Conversely, given the negative impacts of these extreme measures on quality of life and economic activity, these interventions should be planned and evaluated carefully. For these reasons, the model applied here may significantly help to identify determining factors of the success of these external intervention measures, facilitating their control, monitoring, and evaluation.

## Supporting information

Programs (codes, input data and instructions)

Data of stages 1 and 2

## Data Availability

All data used in the present study are available in:
- The article.
- Supplementary Material.
- "Base de Datos COVID-19." http://www.minciencia.gob.cl/covid19 (accessed June 17, 2020).
- "REGION METROPOLITANA DE SANTIAGO. ÍNDICE DE PRIORIDAD SOCIAL DE COMUNAS 2020. Seremi de Desarrollo Social y Familia Metropolitana." http://www.desarrollosocialyfamilia.gob.cl/storage/docs/boletin_interno/INDICE_DE_PRIORIDAD_SOCIAL_2020.pdf (accessed September 7, 2021).
Additionally, data produced in study can be obtained in the article or by using the codes of the supplementary material.

http://www.desarrollosocialyfamilia.gob.cl/storage/docs/boletin_interno/INDICE_DE_PRIORIDAD_SOCIAL_2020.pd

