## Supplementary material for "Modelling Quarantine Effects on SARS-CoV-2 Epidemiological Dynamics in Chilean Communes and their Relationship with the Social Priority Index": Programs (codes, input data and instructions): ajustemodelo1.m

```
function [Ufinalteorico, Qfinalteorico,r, estimates] = ajustemodelo1(Datos, N, alfa0, beta0, gama0, U00, Q00,ultimamedicion, comuna)

%Este programa calcula curva de ajuste y la superpone con los datos experimentales.

% Configuración del ajuste mediante la función fmincon()
A=[];
b=[];
Aeq=[];
beq=[];

lb=[0,0,0,0]; % Mínimos valores de parámetros
ub=[1,1,N,N]; % Máximos valores de parámetros.

start_point= [beta0, gama0, U00, Q00]; % Valores iniciales de arámetros para comenzar el ajuste.

estimates = fmincon(@mifun, start_point, A,b,Aeq,beq,lb,ub);
function sse=mifun(params, file);

% ___________________________________

% Notación para los parámetros de ajuste 
beta = params(1);
gama = params(2);
U0 = params(3);
Q0 = params(4);

alfa=alfa0;

C = Datos(:, 2); % Datos de casos confirmados. 

C0 = C(1);
S0 =N-U0-Q0-C0;

indices=size(Datos);

diastotales=Datos(indices(1,1), 1)-Datos(1, 1)+1;
%__________________________________
% Cálculo del error (sse) obtenido por la desviación entre lo teórico y lo experimental

% Limpiando memoria de variables
clear t
clear y
clear Cteorico
clear Tiempo

% Cálculos de los valores teóricos de C (Casos confirmados teóricos, indicados como Cteorico)
    
   [t,y]= ode45(@campomodelo, [Datos(1, 1) Datos(indices(1,1), 1)], [S0,U0,Q0,C0], [], N, alfa, beta, gama); 

   
    dimensiones = size(y); 
    tiempoenUA=dimensiones(1,1); % Tiempo total, en unidades arbitrarias, del registro de datos. (Es el total de C calculados)
    
for medicion=1:1:indices(1,1) % indices=size(Datos);
    
    i=0;
    for coordenadadetiempo=1:1:tiempoenUA
            
        if t(coordenadadetiempo)<=Datos(medicion,1)
          
        
            Cteorico(medicion)=y(coordenadadetiempo,4);
        elseif i==0
            Cteorico(medicion)=y(coordenadadetiempo,4);
            i=1+1;
                    
        end
    end
    
    ErrorVector1(medicion) = Cteorico(medicion) - C(medicion);
    Tiempo(medicion)=Datos(medicion,1);
  
end

sse = sum(ErrorVector1.^2); % error (función costo, la que debe ser minimizada mediante el ajuste de parámetros)

end

%Este bloque de valores finales de las variables puede ser muy útil en el caso de usar un tiempo final de pandemia. 
Sfinalteorico=y(coordenadadetiempo,1);  
Ufinalteorico=y(coordenadadetiempo,2);
Qfinalteorico=y(coordenadadetiempo,3);
Cfinalteorico=y(coordenadadetiempo,4);


NTotalteoricodelfinal = Sfinalteorico + Ufinalteorico + Qfinalteorico + Cfinalteorico; %Control

Error_porcentual_en_N_calculado = 100*(NTotalteoricodelfinal-N)/N   % Es un control. La variación debe ser por los individuos recuperados.
%__________________________________

if comuna==30 % Se graficará la comuna 30
% A este nivel los parámetros ya se han estimado, minimizando la función error.
% A continuación estos parámtros se ocupan en la generación del gráfico que muestra el ajuste.
% Se grafican la curva teórica y los puntos de datos empíricos.  

plot(Tiempo,C,'ko',t, y(:,4),'--k','MarkerFacecolor','k');

axis([Datos(1, 1),Datos(indices(1,1), 1),0,10000]);
ylabel('Casos confirmados');
xlabel('Días');

end
r= corrcoef(Cteorico,C); %R % (Con r(1,2) el Coeficiente de correlación de Pearson)

end
```
