## Supplementary material for "Modelling Quarantine Effects on SARS-CoV-2 Epidemiological Dynamics in Chilean Communes and their Relationship with the Social Priority Index": Programs (codes, input data and instructions): P2Stage2.m

```
%P2Stage2

clearvars % Limpia la memoria de todas las variables en corridas anteriores del programa.

% Lectura de archivo Excel de datos, cargando los datos a la matriz de datos iniciales Datoscero. 

file=input('Data file (e.g., data_stage2.xlsx): ','s') 

Datoscero = xlsread(file); 

dimensiones = size(Datoscero); % Se obtienen  las dimensiones de la matriz de Datoscero.
    comunas=dimensiones(1,2)-1; % El número de comunas es el el número de columnas menos 1 (pues la primera columna de datos es para el tiempo)
  
file=input('Data file (e.g., U0Q0_for_P2Stage2.xlsx): ','s')     
U0Q0 = xlsread(file); 
dimensiones2 = size(U0Q0);
    
    
    
    for comuna=1:1:comunas %Se hacen los cálculos por comuna.
    comuna %para mostrar en pantalla
    
        %Para obtener número de días de precuarentena 
        
        ultimamedicion=0;
        delta=1;

        for i=2:1:dimensiones(1,1)
        
            if Datoscero(i,comuna+1)==-1
                delta=0;
            end
            
            ultimamedicion=ultimamedicion + delta;        
        end
       
        
    %_______________________________________   
        
    % A continuación se dejan los datos como para ser empleados por la función de ajuste ajustemodelo2().
    
    % Se crea la matriz Datos, la que es solamente para la comuna indicada en el ciclo. Enumera en la primera columna 
    % los días a partir del día 1 y en la segunda columna contiene los casos confirmados iniciales. 
    
 
 
    clearvars Datos;
    
    Datos = [Datoscero(3+ultimamedicion:26+1, 1), Datoscero(3+ultimamedicion:26+1, comuna+1)]; % Hay desplazamiento en 1, pues Datoscero tiene los días a partir de segunda fila,
                                                                   % y los casos confirmados a partir de la segunda columna (y según a qué comuna corresponda la columna).
          
% Población de la comuna.
      N=Datoscero(1, comuna+1);
   
      
% Parámetros iniciales para iniciar después el algoritmo en la función de ajustemodelo2().
alfa0=0.2967; %  para modelo en China sin cuarentena.


beta0=0.385; % 55% sintomático + 70% sensibilidad. Valor inicial tentativo para gama2.
gama0=0.3; % Valor inicial tentativo para gama1.

dimensionesDatos = size(Datos);
medicionesmaximas=dimensionesDatos(1,1);


 U00=U0Q0(2,comuna);

 Q00 =U0Q0(3,comuna);  


%Cálculo del índice de correlación y de los cuatro parámetros de ajuste
[Ufinalteorico, Qfinalteorico,r, estimates]=ajustemodelo2(Datos, N, alfa0, beta0, gama0, U00, Q00,ultimamedicion,comuna); % Se puede consultar el archivo de la función ajustemodelo2(). 

%________________________________________
%Carga en arreglos los resultados de todas las comunas 
comunavector(comuna)=comuna;
Nvector(comuna)=N;
alfavector(comuna)=alfa0;
betavector(comuna) = estimates(1)^2;
gamavector(comuna) = estimates(2)^2;
U0vector(comuna) = estimates(3)^2;
Q0vector(comuna) = estimates(4)^2;
Re_vector(comuna)=alfavector(comuna)/(gamavector(comuna)+1/14);
R2vector(comuna)=r(1,2)^2; % (Coeficiente de determinación para cada comuna)

%________________________________________

    end %cierre de ciclo de cálculos por cada comuna

R2vector % Muestra en pantalla.

% Matriz de resultados para exportar a Excel
Matriz = [comunavector; Nvector; alfavector; U0vector; Q0vector; gamavector; betavector; Re_vector; R2vector]; 

% Creación de archivo Excel con la matriz de resultados
xlswrite('output_P2Stage2.xlsx',Matriz,'Hoja1','B2')
```
