## Supplementary material for "Modelling Quarantine Effects on SARS-CoV-2 Epidemiological Dynamics in Chilean Communes and their Relationship with the Social Priority Index": Programs (codes, input data and instructions): readme.pdf

Before using the programs for the first time, it is recommended to read the following:

1. The MATLAB program has two modes (those that run fine if you keep the files in the same folder and use the MATLAB R2013a version):

Mode 1 (uses fmincon()):
Run P1Stage1 (for stage 1 data) or P1Stage2 (for stage 2 data).

Mode 2 (uses the fminsearch()):
Run P2Stage1 (for stage 1 data) or P2Stage2 (for stage 2 data).

2. The input data are in the files data_stage1.xlsx and data_stage2.xlsx, where -1 has been used to indicate the end and beginning of the data series, respectively. The commune numbers, as well as the data, are from the file "data stage 1 and stage 2 province of Santiago.xlsx", available as a supplementary material.

3. As output you will get a file of type output_PXStageY.xlsx. The file will contain the values of the following parameters: Commune number, N, alpha, U0, Q0, gamma, beta, R and r2, in that order (Note that the notation here is not exactly the same as in the article or in the programs).

4. Additionally, the programs for stage 1 will generate a file of the type U0Q0_for_PXStage2.xlsx, which is required as additional input by the programs corresponding to stage 2.

5. In the execution of the programs, the adjustment for the commune San Ramón (Number 30) is plotted, which can be modified in the codes.
